# Measures of retention in HIV care: A protocol for a mixed methods study

**DOI:** 10.1101/2023.11.10.23298385

**Authors:** Nadia Rehman, Michael Cristian Garcia, Aaron Jones, Jinhui Ma, Dominik Mertz, Lawrence Mbuagbaw

**Author notes:** **Corresponding Author:** (LM). These authors contributed equally to this work (Michael Cristian Garcia, Aaron Jones, Jinhui Ma, Dominik Mertz).

## Abstract

**Introduction:** Retention in HIV care is necessary to achieve adherence to antiretroviral therapy, viral load suppression, and optimal health outcomes. There is no standard definition for retention in HIV care, which compromises consistent and reliable reporting and comparison of retention across facilities, jurisdictions, and studies.

**Objective:** The objective of this study is to explore how stakeholders involved in HIV care define retention in HIV care and their preferences on measuring retention.

**Methods:** We will use an exploratory sequential mixed methods design involving HIV stakeholder groups such as people living with HIV, people involved in providing care for PLHIV, and people involved in decision-making about PLHIV. In the qualitative phase of the study, we with conduct 20-25 in-depth interviews to collect perspectives of HIV stakeholders on using their preferred retention measures. The interview guide has being provided as an online Supplementary Appendix 1.The findings from the qualitative phase will inform the development of survey items for the quantitative phase. Survey participants (n=385) will be invited to rate the importance of each approach to measuring retention on a seven-point Likert scale. We will merge the findings from the qualitative and quantitative findings phase to inform a consensus-building framework for a standard definition of retention in care.

**Ethical Issues and Dissemination:** This study has received ethics approval from the Hamilton Integrated Research Ethics Board. The findings will be disseminated through peer-reviewed publications, conference presentations, and among stakeholder groups.

**Limitations:** 1. This study has limitation, we won’t be able to arrive at a standard definition, a Delphi technique amongst the stakeholders will be utilized using the framework to reach a consensus globally accepted definition.

## Introduction

Globally, 38.4 million people are living with HIV, with 1.5 million new infections acquired in only in 2021[1]. In 2015, the United Nations set the Sustainable Development Goals (SDGs) to end the HIV epidemic by 2030 [2]. To reach the SDGs, efforts have been made to make free antiretroviral therapy (ART), and quality care accessible for people living with HIV (PLHIV) on the global scale [3]. However, we are far from achieving the SDGs, and even though HIV care is available, PLHIV are not retained in care owing to societal inequalities, lack of uniform policies, and HIV-related stigma [4].

To meet the SDGs, it is crucial to retain patients in every stage of the HIV care continuum. Even though ART helps prolong and improve the quality of life of PLHIV, poor adherence to ART can lead to virological failure, progression to AIDS, and the development of resistant strains [5]. Retention in care is essential for ensuring consistent access to ART, monitoring of health outcomes, lowering mortality rates, and reducing new HIV infections [6–8]. Adequate retention in care is known to be cost-effective and maximize patient outcomes [9].

The public and individual health implications of retention in HIV care, necessitates an accurate estimate of retention in care. The lack of a standard and globally accepted definition of retention has been highlighted in the literature many times [10–16]. Consensus on a standard definition is compromised by the need for longitudinal assessment and the various socio-economic factors predicting retention in PLHIV [10, 16, 17]. In our recently published review of definitions of retention, we identified that trialists have used heterogeneous definitions of retention across studies [18]. This limits our ability to aggregate data for meta-analysis and to compare findings across studies. A standard definition must be in place to report and compare long-term retention across jurisdictions, settings, and studies [12]. A reliable and consistent measure of retention will be a starting point to identify real-world gaps, evaluate program performance, make policies, and implement them to ensure equitable healthcare access [19].

A standard definition must include clinically relevant features and be adequately stable to predict long-term clinical outcomes. The definition should be easy to use, record and report across different settings, healthcare systems, and populations. Arriving at a consensus for a standard definition will require views and perspectives from stakeholders involved in HIV care, including PLHIV [18].

The purpose of this study is to develop a standard definition of retention in care with consensus from global stakeholders in HIV care. For this study, we will define HIV stakeholders in three main categories based on the evidence to inform policy (policymakers), health research (researchers and funding bodies), and practice (HIV clinicians, clinical guideline developers, public health professionals, and PLHIV) nationally and internationally [20]. The following research questions guide our study.

### Qualitative research questions

- What are the various perspectives of HIV stakeholders engaged in HIV care at national and international levels on measuring retention in HIV care?
- How do the categories mentioned by the HIV stakeholders help to explain the need for a standard measure of retention in HIV care?

### Quantitative research questions

- What components used to define retention are most important to HIV care stakeholders in terms of ease to use, recording, and comparison across different settings, jurisdictions, and studies?

### Mixed method research questions

- To what extent and in what ways do qualitative interviews with stakeholders involved in HIV care services contribute to a comprehensive understanding of the various components used to define retention in care?
- Are the views of HIV stakeholders about measuring retention in their specific context generalizable to HIV stakeholders in a broader context?

## Methods

We will use the pragmatism paradigm as the epistemological framework for our research. Pragmatism is an appropriate approach for our research as we will be examining diverse and complex contexts [21]. Pragmatism will provide us with the flexibility to build variations into techniques and analytic processes, resulting in the production of actionable knowledge [22].

### Study design

We will conduct an exploratory sequential mixed methods study (QUAL → quan) [23]. The mixed method approach will comprise two phases. In the qualitative phase, we will conduct in-depth interviews building upon retention in care definitions identified in a prior review [18]. In the qualitative phase, we will explore participants’ views on measuring retention with the intent of using this information to develop a survey. In the quantitative phase of the study, the key concepts identified from the qualitative data will be operationalized as quantitative items in a web-based survey. This survey will enable us to generalize the qualitative findings to a larger heterogeneous sample. To achieve complete and comprehensive answers to our research question, qualitative and quantitative findings will be merged for analysis and interpretation, reporting and development of the framework for consensus of the standard definition [23, 24].

### Rationale for design

A mixed-method approach will address the complex and broader question of developing a standard and optimal measure of retention [25]. A mixed-method approach will enable us to use two sampling techniques [26].

Purposeful and probabilistic techniques will ensure data collection in breadth and depth [27]. A qualitative phase of individual interviews will help contextualize and elaborate findings from different settings, populations, and professional backgrounds [28]. The quantitative data will inform us of the magnitude of the relevance of the retention measures [21]. This approach will generate findings that would not otherwise be obtainable when using only qualitative or quantitative methods alone. This mixed-method approach will increase the validity of our interpretations and generate reliable new knowledge [29].

### Study Timeline

The participants recruitment for the qualitative phase of the study willl begin in November 2023 and will continue until March 2024. The qualitative synthesis will occur between April 2024 and June 2024. From the qualitative findings we will develop the survey.The survey will be disseminated from August 2024 till November 2024. The quantitative findings and the integration of the qualitative and quantitative findings will be finalized from December 2024 till February 2025.

### Sampling and sample size

#### Qualitative phase

The qualitative phase of our study will draw on the principles of qualitative description methodology. This approach is suitable for mixed method study to address clinical questions. This approach is well suited here as this approach will enable us to gain perspectives from participants to understand and describe the phenomenon in terms of literal descriptions and dwell on practical relevance of the information for health care users [30].

The participants recruitment will begin in November 2023, and will continue until March 2024. Participants will be selected if they meet the following inclusion criteria: 1) able to understand both written and spoken English; 2) a minimum age of 18 years of age; 3) are willing to participate in a computer-based online interview about personal experiences with retention in care; and 3) can provide signed informed consent for study participation. Participants will be excluded if they do not participate in the interview or request that their data be withdrawn.

We will employ a combination of purposeful sampling techniques, including criterion-based, maximum variation, and snowballing sampling techniques, to seek an equally representative sample in terms of the HIV stakeholders groups (policy, research, practice) and across all WHO geographical regions (African Region, Region of the Americas, South-East Asia Region, European Region, Eastern Mediterranean Region, and Western Pacific Region) [31].

Based on sample size recommendations to reach theoretical sampling, we anticipate performing 25-30 interviews, with 4-5 from each representative group. This sample size seems appropriate to capture information-rich data. This estimation of interview participants is based on a study involving key stakeholders’ perspectives and experiences on defining and identifying health research gaps [20].

#### Quantitative phase

We will use the email listservs of organizations working in HIV care, such as the Ontario HIV Treatment Network (OHTN), World Health Organization (WHO) and Joint United Nations Programme on HIV/AIDS (UNAIDS), Global Fund, International AIDS Society (IAS), Kaiser Family Foundation (KFF), Canadian Foundation for AIDS Research (CANFAR) and regional WHO groups. Researchers will be identified from publications on retention in care in the CASCADE database (a database of randomized trials on the HIV care cascade) [32]. We will also reach out to existing collaborations and professional associations such as the International AIDS Society (IAS), the Canadian Association for HIV Research (CAHR), and the International Association of Providers of AIDS Care (IAPAC).

To further supplement the recruitment process, we will publicize our study on social media platforms like Twitter and LinkedIn. Patients will be recruited and identified from online patient support groups and organizations serving those patients, such as REALIZE, POZ Community Forum [31]. We will submit an amendment to HiREB regarding any changes in recruitment methods based on guidance of the organizations.

We will disseminate the survey online in the stakeholders’ groups. If response rates are lower than our calculated sample size, we will use a snowballing approach to include responses from participants interested in this topic [33].

We will use a proportional quota sampling method to achieve a diverse and representative sample of participants. We will divide the sample into our pre-defined three strata (policy, health research, and practice) so every HIV stakeholder group is equally represented.[34] People who belong to more than one group will be asked to declare the group they primarily identify with. We will obtain a pre-determined sample population of 385 based on the formula for proportions for a survey, which is the maximum sample size required to estimate a population proportion of 50% with a 95% level of confidence and a 0.5% margin of error. These computations are performed with WINPEPI.[35]

### Data collection

#### Qualitative phase

We will conduct in-depth semi-structured interviews following a piloted interview guide. A copy of the interview guide is provided in the Supplementary Fil1. The interviews will be conducted over Zoom (a videoconferencing platform with real-time messaging and content sharing). The interview time will be booked according to the participants’ preferrence and will last approximately one hour.

At the beginning of the session, the trained interviewers will cover the study’s purpose, consent, and ethical issues, and establish rapport with the interviewees.

All the interviews will be recorded and transcribed verbatim by using a transcription service. About 10% of the transcripts will be compared by two authors (NR, CG) to the data transcribed by the transcription service for congruence and inconsistencies. Interview data will be anonymized and stored on secure and password-protected servers at McMaster University.

#### Quantitative phase

This survey will be designed according to expert recommendations for electronic surveys [36].The web-based quantitative survey for participants will be conducted on the Research Electronic Data Capture (REDCap) tool, a secure, web-based application hosted and stored on a secure server at McMaster University [37]. The survey will be piloted before deployment. Participants will be presented with an overview of the study, the study’s purpose, the privacy and confidentiality of their data, and the time required to complete the survey. Participants will be invited to complete a consent form. The survey will be a self-administered, structured questionnaire in English.

The survey will consist of three parts: demographic information, professional information and questions related to measures of retention in HIV care. The themes identified in the qualitative interviews will serve as the survey items on the measures of retention. The participants will rate them on a 7-point Likert scale [38].

To ensure an adequate response rate, email addresses will be updated, and at least two reminder emails will be sent to potential participants.

### Analysis

#### Qualitative phase

We will conduct a qualitative content analysis [39]. Transcriptions will be analyzed using content analysis techniques supported NVivo 10.

Both deductive and inductive content analysis procedures will be used in the following steps: (1) the initial coding framework for the deductive analysis will be informed by definitions of retention identified in our previous work [40]; inductive codes will be derived from the data by iterative readings of the interview transcripts. Similar codes will be put into categories according to their similarities and differences. The responses along with the number of participants in each category will be counted. The various categories will be organized in a table to create a visual and contextual interpretation [30].

After every seven to eight interviews, the two coders will meet to compare categories and arrive at an agreed-upon set of categories. Disagreement will be resolved by discussion to improve reliability between the two coders.

#### Quantitative phase

We will compute the mean (standard deviation) or frequency (percent) based on the survey responses as appropriate for the data.

To determine the level of importance that participants allocate to each item we will compute the mean (SD) or median (quartiles). Only items with a mean of 4 or more out of seven will be considered as important. The mean will inform us which approaches to measuring retention are preferred by the participants. Results will be aggregated by stakeholders groups.

The results will be analyzed using IBM SPSS statistical software [41]. An outline of the study is shown in Figure 1. Study diagram of an exploratory sequential study with survey development on measures of retention in HIV.

### Data integration

Data integration will occur at every stage of the study to answer our research question in depth and breadth:(1) connecting both the data through a sampling frame: participants are selected from the population of participants who responded to the survey (2) Typology development (building): the survey will be built on the themes identified in the qualitative data [21].

Three approaches will be used to integrate our qualitative and quantitative findings at the interpretation and reporting level: First we will transform the qualitative data into quantitative and the data will be displayed in a table [42]. This method will allow to inform the analysis of qualitative data to inform the quantitative finding [26].

Second, we will use joint displays to visually integrate and report the findings of both phases. The joint displays will allow the visual presentation of the data in matrix. The first joint display will show the link between the qualitative findings and how these findings tailored the methodology and development of the quantitative phase, including the survey items [43]. The second joint display will depict how the quantitative finding reflects and generalizes the qualitative context. The joint display tables will show the results together organized by common categories so that a comparison can be made to come to a final conclusion, i.e., ‘meta-inferences’[44].

Third, we will use contiguous narrative data integration and reporting approach, presenting qualitative and quantitative findings separately in the same paper.

### Validation checks

We will use the Lincoln and Guba theoretical framework for mixed-methods studies to include parallel quantitative and qualitative criteria for criteria appraisal to promote the rigor and trustworthiness of our procedures and findings strategies [25, 45]. In the qualitative strand, we will email the transcripts to the partcipants for member checking to ensure study credibility. We will enhance consistency in the qualitative phase by peer debriefing and involving other authors in the transcription and analysis procedure to identify any overlooked biases [27]. We will keep an audit trail of the decisions made in all the study steps [46]. We will maintain a reflexive journal to explore the researchers’ subjectivity [47, 48]. For the quantitative phase, we will increase the accuracy and reliability of our findings by keeping the questionnaire clear and easy to read by pilot testing the questionnaire [26, 49].

We will check the internal consistency of survey responses with Cronbach Alpha coefficient (higher than 0.8), indicating high consistency across survey items [25].

## Discussion

### Overview

This study will provide important insights into HIV stakeholders’ views and opinions on measuring retention best and overcoming the variability in the measures. This knowledge will add to the improvement of research, policy, and services and contribute to improvements in the quality of care for PLHIV.

This study is a follow-up study to a prior project. Previously we conducted a SWAR to identify definitions of retention used in RCTs. This review showed a wide heterogeneity in the definitions of retention [18]. We anticipate that this mixed-method approach will provide the breadth and depth of the topic and will advance efforts towards a standard measure of retention in care.

### Ethics

This research proposal has received ethics approval from the Hamilton Integrated Research Ethics Board ethics committee for approval (HiREB). Participants’ confidentiality and anonymity will be respected at all times. Written informed consent will be obtained from all participants, however the participants can withdraw at any time. The transcripts of all interviews will be de-identified before analysis. No personally identifying data will be collected during the surveys. All recordings,transcripts and survey data will be stored in password-protected and encrypted servers at McMaster University. All data will be destroyed after ten years.

### Dissemination

Our study will be disseminated via three peer-reviewed publications (i.e., the study protocol, a qualitative study, and a mixed-method paper)and as part of a doctoral thesis. We will also disseminate this at relevant conferences and other stakeholder gatherings.

## Data Availability

For maintaining the privacy of the participants only deidentified data will be collected, and per HiREB that data won't be shared to maintain the confidentiality of the participants.

## Acknowledgements

All the researchers (L.M., N.R., M.G.,J.M., D.M., A,J.) involved will be granted authorship, because of their substantial contributions to the work.

## Supporting information

**S 1 Fig.** Study diagram of an exploratory sequential study with survey development on measures of retention in HIV.

**S2 Appendix.** Interview guide for the qualitative phase of the study.

